# Hypertrophic cardiomyopathy: a genome wide association meta-analysis and polygenic risk score

**DOI:** 10.64898/2026.05.05.26352435

**Authors:** Luis R Lopes, Nay Aung, Stefan van Duijvenboden, Hannah L Nicholls, Richard Burns, Joanna Jager, Massimiliano Lorenzini, Mohammed M Akhtar, Alexandros Protonotarios, Cayetana Barbeito-Caamaño, Jose María Larrañaga-Moreira, Roberto Barriales-Villa, Kayesha Coley, Chiara Batini, Gerald Sze, Martin D Tobin, Catherine John, Steffen E Petersen, Petros Syrris, Patricia B Munroe, Perry M Elliott

**Author notes:** **Corresponding author:** Luis R. Lopes, Barts Heart Centre, St. Bartholomew’s Hospital, West Smithfield, London EC1A 7BE, United Kingdom. these authors contributed equally to the manuscript and are joint first authors (Luis R Lopes, Nay Aung). these authors contributed equally to the manuscript and are joint last authors (Patricia B Munroe, Perry M Elliott).

## Abstract

**Background:** Hypertrophic cardiomyopathy (HCM) is a heritable trait with marked variability in expression and outcomes. Our aims were to discover new genetic loci associated with HCM and to test the effect of a new polygenic risk score (PRS) on incidence, phenotype and outcomes, stratified by sarcomere genotype status.

**Methods:** A discovery genome-wide association study (GWAS) was performed on 2,284 HCM cases and 4,525 controls. Two fixed-effects meta-analyses combined our discovery GWAS with single-trait and multi-trait results from a published study. Discovered loci underwent comprehensive bioinformatic analysis including functional and druggability annotations. A PRS using loci from the two meta-analyses was evaluated for association with: HCM diagnosis in 411,213 individuals from UK Biobank (UKBB); imaging phenotypes in individuals without HCM; a composite endpoint (including all-cause mortality and transplantation) and sudden cardiac death (SCD) in 1,756 HCM cases. PRS analyses were stratified by sarcomere genotype status.

**Results:** Three loci were found in the discovery GWAS (*BAG3, FHOD3* and novel locus *PPP1R3A*). In the meta-analyses, 70 unique loci were identified, four novel (*MYPN, YWHAE, NOS1AP* and *OBSCN*). Bioinformatic analyses identified NOS1AP as a candidate HCM gene. A new PRS was significantly associated with HCM diagnosis (hazard ratio [HR] 3.19, 95% CI:2.46–4.14, for top 5% vs lower 95%; HR 1.88, 95% CI:1.72–2.06, per SD increase). Significant associations were found between PRS and greater left ventricular (LV) wall thickness and higher LV ejection fraction in UKBB participants without HCM. Sarcomere-negative HCM cases in the top 20% of the PRS distribution had an increased risk of SCD (HR 2.72, 95% CI:1.03–7.17).

**Conclusions:** We report novel HCM loci. A new PRS predicted the risk of HCM development and associated imaging characteristics in the UKBB and outcomes in an HCM cohort.

## INTRODUCTION

Hypertrophic cardiomyopathy (HCM) is defined by a single phenotype trait, left ventricular wall thickness (MLVWT) ^1^, and while often inherited in an autosomal dominant pattern caused by pathogenic variation in cardiac sarcomere genes, more than half of all cases remain genetically elusive ^2^. Familial linkage analyses and, more recently, whole-exome sequencing (WES) have revealed new causal genes, each of which account for a small proportion (1-2%) of cases ^3,4, 5, 6^.

The role of complex genetic inheritance in HCM has been explored. Two genome-wide association studies (GWAS) ^7,8^ have supported a polygenic influence on the HCM phenotype that was most marked in patients without dominant sarcomere gene variants ^7^, findings that were corroborated in a recent study ^9^ . A PRS derived from one of the GWAS ^7^ was shown to be associated with increased susceptibility for HCM in the population over rare variant and clinical risk factors ^10^. A PRS ^9^ was reported to enable risk stratification for developing HCM and adverse outcomes among relatives and the risk of events in HCM cases. Additionally, a case-control GWAS involving 5,900 HCM cases and 68,359 controls, alongside a multi-trait analysis of GWAS (MTAG) incorporating 36,083 UK Biobank participants with cardiovascular magnetic resonance imaging, identified 70 loci (including 50 novel loci) for HCM and 62 loci associated with relevant left ventricular traits, of which 20 were novel ^11^.

Our study aimed to discover novel HCM loci by meta-analysing an independent HCM case-control dataset with the largest published HCM GWAS, and to develop a new PRS in order to better predict disease incidence and patient outcomes, stratified by the presence of causal sarcomere gene variants.

## METHODS

A flowchart for quality control/sample selection is shown in **Supplementary Figure 1** and a flowchart for the whole study design in **Figure 1**.

**Figure 1.**
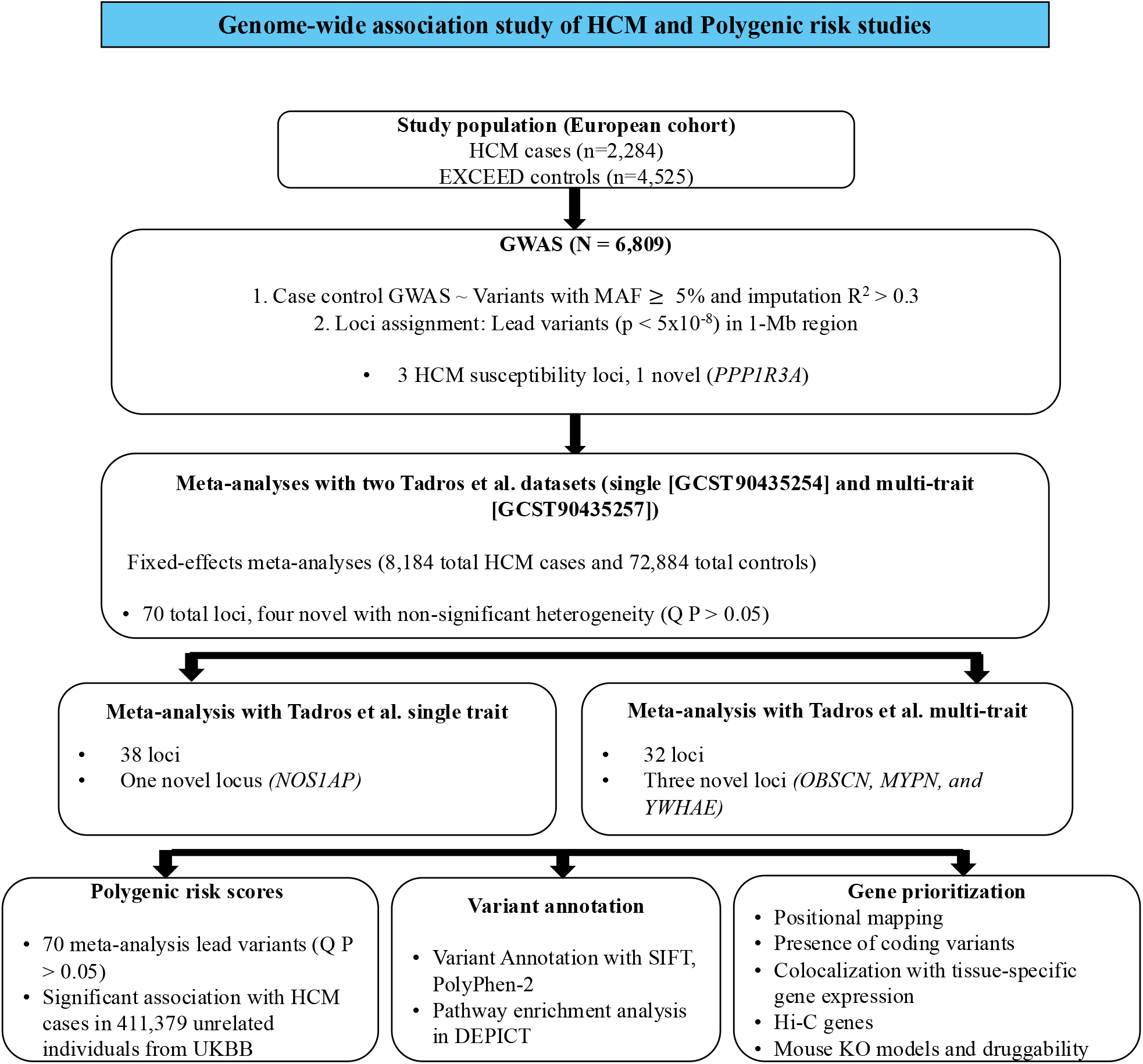
Study flowchart.

### Study population, targeted panel sequencing and exome sequencing

The patient cohort comprised individuals with HCM referred to the Inherited Cardiovascular Disease unit at St. Bartholomew’s Hospital, London, UK (2015-present), the Inherited Cardiovascular Disease Unit at The Heart Hospital, University College London Hospitals (UCLH), London, UK (before 2015) and the Unidad de Cardiopatías Familiares of Complexo Hospitalario Universitario A Coruña, Spain. All patients gave written informed consent for genetic testing, and the study was approved by the regional ethics committees (London: 15/LO/0549; Coruna: 2021/182). Clinical parameters were recorded using previously described methods and were stored in a dedicated database ^12^. HCM was defined by the presence of a maximal left ventricular wall thickness ≥ 15mm in probands or ≥ 13mm in relatives ^1^. Patients with previously confirmed HCM phenocopies were excluded from the study. The samples used in this study were collected from 2011 to 2019 in London and 1991 to 2021 in A Coruna.

In more detail, the London study cohort comprised 874 consecutive HCM probands screened with targeted next-generation sequencing (NGS) for the whole-genomic region of 41 cardiovascular genes ^13^ and 770 consecutive index cases tested using whole-exome sequencing (WES) ^3^; the remaining consecutive probands, including those from Spain, were tested with clinically requested targeted panels that always comprised established causal sarcomere genes and phenocopies.

DNA extraction, library preparation, whole-exome sequencing, variant calling and annotation were performed as described previously ^3^. Rare variants were classified as pathogenic, likely pathogenic, unknown significance, or likely benign/benign using the current criteria of the American College of Medical Genetics and Genomics ^14^. Patients with ≥1 pathogenic or likely pathogenic variant were designated as sarcomere-positives, while those with no pathogenic or likely pathogenic variants were designated as sarcomere-negatives.

Detailed descriptions and definitions of the phenotype parameters and outcomes analysed are reported in **Supplementary Methods**.

### GWAS sample – genotyping, quality control and imputation

The HCM samples from the two centres (n=3,024) were genotyped at University College London (UCL) Genomics unit using the Infinium Global Screening Array-24SA v3.0 with a standard methodology (https://emea.illumina.com/products/by-type/microarray-kits/infinium-global-screening.html). Ancestry estimation was performed using an ancestry identification tool from GenoPred that infers the genetic ancestry of a target sample based on a principal component (PC) elastic net model ^15^ projected onto the 1000 genome project reference panel ^16^. After removing individuals with high genotype missing rate (>5%), extreme heterozygosity exceeding three standard deviations from the mean, duplicate participant data and inferred sex discordance, we included 2,284 HCM cases with European ancestry in subsequent analyses.

The controls for this study were derived from the EXCEED (Extended Cohort for E-health, Environment and DNA) study ^17^. EXCEED is a longitudinal population-based cohort which facilitates investigation of genetic, environmental and lifestyle-related determinants of a broad range of diseases and of multimorbidity through data collected at baseline, via electronic healthcare records linkage and recurrent self-completion questionnaires. Launched in 2013, EXCEED initially recruited males and females, aged 30-69 and living in Leicestershire and Rutland in the UK via smoking clinics and GP practices. In May 2020, recruitment opened to anyone over the age of 18 years, living in either the East or West Midlands UK regions and registration and consent are now completed online via the EXCEED website. To date, some 11,200 participants have consented to follow their electronic health records for up to 25 years, storage and analysis of their DNA sample and to be contacted for ‘recall-by-genotype’ and ‘recall-by-phenotype’ sub-studies. Genome-wide genotyping for EXCEED participants was performed using the Affymetrix UK Biobank Axiom Array. EXCEED participants of European ancestry free from known HCM and other cardiovascular conditions were identified using a predefined set of 370 clinical Read codes, relating to cardiac arrest, heart failure, cardiomegaly, valve diseases, glycogen storage disorders, amyloidosis, and muscle disorders. This resulted in 4,525 individuals being selected as controls.

Cases and controls were genotyped on different arrays (cases on the Illumina Global Screening Array and controls on the Affymetrix UK Biobank Axiom Array). To reduce the possibility of platform-related batch effects, we restricted analysis to overlapping high quality directly genotyped variants (excluding variants with missingness > 1%, MAF < 1%, and Hardy Weinberg P values of 1x10^-15^) in both datasets, and then performed harmonisation and QC using the Wrayner tool (v4.3.0) (https://www.well.ox.ac.uk/∼wrayner/tools/) which provided strand alignment check and removal of palindromic (ambiguous) SNPs. Joint imputation was performed on the merged genotype data using the Michigan Imputation server and the Haplotype Reference Consortium (HRC) reference panel ^18,19^.

### GWAS – case-control discovery analysis

The association analysis was performed using REGENIE (v3.0) which uses a two-step approach ^20^. In Step 1, a whole genome regression model was fit on directly genotyped variants (MAF⍰>⍰0.05; genotype missing rate < 1%, HWE test P-value > 1⍰×⍰10^−9^) with leave-one-variant-out cross validation to avoid overfitting. In Step 2, predictions from Step 1 were used when testing the phenotype against the imputed variants, using the leave-one-chromosome-out scheme with the fast Firth correction as a fallback for p-values below 0.01. The first 10 ancestry-informative principal components (PCs), derived from the high-quality variants selected for step 1 using PLINK (v2.00a2LM) were included as covariates. Genome-wide significant (GWS) threshold was set at P⍰<⍰5⍰×⍰10^−8^, after excluding variants with MAF <5% and imputation R2 <= 0.3. We defined genome-wide significant loci by assigning lead variants (P < 5 x 10^-8^) to 1-Mb loci blocks, grouping together variants located within the same 1-Mb window and in LD (r^2^ > 0.4). We applied a more stringent MAF threshold to account for the relatively modest sample size of the discovery case-control cohort. Loci were annotated as novel via cross-referencing Tadros et al. ^8^, Harper et al. ^7^, and Tadros et al. ^11^ as well as programmatically searching the GWAS catalog for any HCM associated loci within 1Mb (accession date: 22/05/2025). We also performed a sensitivity analysis, restricting the HCM cases to the London cohort (n= 1,539 cases) against the 4,525 individuals in the EXCEED control cohort, and conducted a GWAS to re-evaluate the case-control associations of our lead variants.

### Meta-analysis

We conducted two meta-analyses integrating our GWAS summary statistics with recently published GWAS data from the Tadros et al study which performed single-trait and multi-trait HCM GWAS ^11^. Specifically, we meta-analysed our GWAS discovery summary statistics data with: (i) HCM case-control single-trait summary statistics from Tadros et al. (GWAS Catalog ID: GCST90435254), which included 5,900 HCM cases and 68,359 controls, and separately with their (ii) HCM MTAG analysis which leveraged the GWAS of ten left ventricular traits from the same study (GWAS Catalog ID: GCST90435257). The three studies were harmonised, accounting for allele flipping and using build GRCh37 in R (v4.1.1). Meta-analysis was performed using fixed-effects inverse-variance weighted meta-analysis via GWAMA (Genome-Wide Association Meta-Analysis) software (v2.2.2). We primarily considered significant loci (P < 5x10^-8^) with evidence of low heterogeneity (Cochran’s Q P > 0.05) in downstream analyses. For lead SNPs (single nucleotide polymorphisms) identified by both meta-analyses, we used the effect sizes of the lead variants from the single-trait meta-analysis.

### Bioinformatic analyses and gene prioritisation for the GWAS generated variants

#### Variant annotation

Lead variants and their proxies (r^2^ <0.01) were filtered to 99% credible sets using approximate Bayes factors as previously described by Wakefield. We used Variant Effect Predictor (VEP) ^21^ to describe the type, consequence and predicted function based on SIFT and PolyPhen-2 of all variants in high linkage disequilibrium (LD) (r^2^ ≥ 0.8) with the lead variants from both the discovery and meta-analysis. Non-synonymous variants were considered ‘damaging’ via CADD > 20 or RegulomeDB rank <= 1.

#### Long-range chromatin interaction (Hi-C) analysis

To identify distal candidate genes, we explored chromatin interaction (promotor capture Hi-C) ^22^ data for all variants in LD (r^2^>0.8) with our sentinel SNPs with regulatory potential (RegulomeDB score ≤ 5 and RegulomeDB probability > 0.75) ^23^. We identified genes whose promoter regions form significant chromatin interactions in cardiac atria, ventricles, and aorta tissues. Interactors with the highest regulatory potential were used to annotate the loci.

#### Colocalisation with expression data

We conducted colocalization analysis of our GWAS loci and cis-eQTL signals from the heart and arterial tissues (left atrial appendage and left ventricle, artery aorta, and artery coronary) in the GTEx database (v8) ^24^ using the COLOC software package in R ^25^.

#### Pleiotropy analyses

We queried our HCM case-control GWAS lead variants and their proxies at linkage disequilibrium (LD) r^2^ ≥ 0.8 in the GWAS Catalog ^26^ (queried on 30^th^ April 2025) to investigate pleiotropic associations.

#### Enrichment

We performed Data-driven Expression Prioritized Integration for Complex Traits (DEPICT) ^27^ analysis including all SNPs achieving a suggestive statistical significance (P⍰<⍰5⍰×⍰10^−5^) to detect gene sets and tissues that are enriched for HCM loci. We also performed enrichment analysis utilizing the International Mouse Phenotyping Consortium (IMPC) mouse model phenotypes, identifying significantly enriched knockout phenotypes via adjusted p-values using false discovery rate<0.05 using the Benjamini-Hochberg method.

#### Gene Prioritisation

All genes located within a 10kb window around the identified loci from both discovery and meta-analysis were annotated. Initially, gene prioritization was conducted using PoPS (Polygenic Priority Score), which integrates gene-level association statistics from GWAS summary data and assesses polygenic enrichment across multiple biological features, including cell-type-specific gene expression, biological pathways, and protein-protein interactions. To strengthen the prioritization process, we incorporated additional bioinformatic resources and systematic scoring. Each gene was evaluated and assigned a score from 1 to 3, where 3 represented the highest level of supporting evidence and 1 the lowest. The scoring framework considered multiple lines of evidence, including 1) PoPS, 2) OMIM or ClinGen annotations for cardiovascular disease, mouse model cardiovascular phenotypes, and 3) enrichment in GTEx cardiovascular tissues or the presence of Hi-C data indicating relevant chromatin interactions. Equal weighting was applied across these three criteria, with genes receiving a score of 3 if they were supported by all three annotation groups, while those with only one supporting annotation were assigned a score of 1. Prioritized genes were annotated with gene-drug interactions and drug warnings from Open Targets (v24.03), and gene-drug interactions and druggability categories from DGIdb (June 2023 release).

### Association between Polygenic Risk Score and HCM in the UKBB

A weighted polygenic risk score (PRS) was constructed using the lead variants selected per locus from unique genome-wide significant loci (P < 5x10^-8^ and non-significant heterogeneity Q P > 0.5) from two (single-trait and multi-trait) GWAS metanalyses (see Meta-analysis methods above). The lead variants were selected for each locus based on having the most significant p-value within that locus group.

We evaluated our PRS association with HCM diagnosis in 411,213 unrelated individuals from UKBB. In each individual, the cumulative genetic effect of selected lead variants was calculated using the allelic scoring function in PLINK, where the relative weight assigned to each SNP was the effect size determined in the GWAS. The PRS was residualised for the first 5 principal components (PCs) of genetic ancestry and standardised to mean 0 and standard deviation 1. Multivariable-adjusted Cox proportional hazards regression models were used to investigate the association between PRS and HCM. Individuals were censored at death or the end-of follow-up. We considered PRS as a continuous measure (units: per SD increase), but as the association might be non-linear, we also evaluated the HCM risk associated with the top 5% PRS compared to the bottom 95%. We included age and sex as potential confounders. In addition, to address potential confounding by hypertension, we further adjusted for hypertension status (yes/no). All HCM cases (incident and prevalent) were identified via linkage to Hospital Episode Statistics (HES; hospital diagnoses from the UK National Health Service) and the UK death registry using ICD10 codes I42.1 and I42.2. Hypertensive individuals were identified using UKBB data-field 20002 (Description: non-cancer illness code, self-reported) mentioning “hypertension” or “essential hypertension” and data-field 6150 (Description: Vascular/heart problems diagnosed by doctor) mentioning “high blood pressure”; these data are included in the self-reported assessment at recruitment.

We also performed PRS-imaging association analyses in the UKBB population with available cardiovascular magnetic resonance (CMR) data (n=56,945). This analysis was restricted to individuals without a recorded diagnosis of HCM. Linear regression models were used to test the association between the PRS and three quantitative imaging traits: left ventricular ejection fraction (LVEF), maximum left ventricular wall thickness (MLVWT), and left atrium (LA) diameter. All models were adjusted for age, sex, height, weight, genotyping array, imaging centre, and population principal components (PC1–PC10). In addition to per-standard-deviation effects, we compared individuals in the top 5% of the PRS distribution with the lower 95% to explore potential nonlinear effects. The published Tadros et al. GWAS incorporated 36,083 UKBB participants with CMR data in the MTAG analysis. Consequently, the meta-analysis summary statistics used to construct the PRS include UKBB samples that partially overlap with the cohort used for PRS evaluation.

### Association of PRS with phenotype and outcomes in HCM cases

In 1,756 HCM cases from our discovery analysis with available imaging and outcome data, we tested the association between the PRS and intermediate imaging phenotypes (LA diameter, MLVWT and LVEF) by linear regression. In the same cohort, we evaluated the influence of PRS on incident outcomes which included (i) a composite endpoint of all-cause mortality, cardiac transplant and sudden cardiac death (SCD) or equivalent (defined as aborted SCD or appropriate shock from implantable cardioverter defibrillator), and (ii) an endpoint of SCD or equivalents alone, using Cox proportional hazards analysis. Using the same framework applied in the UKBB PRS analysis, we tested the PRS in HCM cases as continuous measure (per SD increase) and a dichotomised version. For the latter, due to the smaller sample size, we compared the top 20% vs the bottom 80%. Patients who experienced an event prior to the recruitment were excluded. Patients who did not experience an event were censored at death or the end of the study period. Additionally, we stratified all these analyses by the presence of a rare causal variant in an established sarcomere gene (sarcomere-positive and sarcomere negative cases).

## RESULTS

The discovery GWAS comprised 6,809 European individuals of whom 2,284 were HCM cases and 4,525 were EXCEED study controls. The mean age of the HCM cohort was 54.0 (16.8) years and 1,485 (65%) were men. Among cases with HCM with available outcome data who were previously genotyped (1,756 from both centres), 38.2% carried likely pathogenic or pathogenic variants in known sarcomeric genes (sarcomere-positive) and the remaining cases were classified as sarcomere-negative. EXCEED participants’ characteristics are presented in **Supplementary Table 1**.

### Genomic loci associated with HCM in the discovery GWAS

The discovery case-control GWAS identified three HCM susceptibility loci (*BAG3, FHOD3*, and *PPP1R3A*) at P < 5 x 10^-8^ (**Table 1 and Figure 2**), with *PPP1R3A* <u>being a novel HCM locus</u> (**Table 1**). There was no significant genomic inflation (λ = 1.082, LD score regression intercept = 1.076) (**Supplementary Figure 2**). The regional plots of the loci are depicted in **Supplementary Figure 3**. All three loci had concordant statistical significance on sensitivity analysis when performing the GWAS using only the London HCM cases with the EXCEED controls (**Supplementary Table 2**).

**Table 1.** Discovery HCM GWAS loci.

| Locus number | Locus | rsid | SNP | EA | NEA | EAF | OR (95% CI) | P |
| --- | --- | --- | --- | --- | --- | --- | --- | --- |
| 1 | *PPP1R3A* | rs12536048 | 7:113674529:C:T | T | C | 0.71 | 0.71 (0.63 - 0.79) | 2.75E-10 |
| 2 | *BAG3* * | rs72842207 | 10:119674163:T:C | C | T | 0.81 | 0.69 (0.61 - 0.78) | 5.23E-09 |
| 3 | *FHOD3* * | rs1893433 | 18:36652989:C:T | T | C | 0.80 | 0.71 (0.63 - 0.80) | 4.43E-08 |

SNP, single nucleotide polymorphism; EA, effect allele; NEA, non-effect allele; EAF, effect allele frequency; OR, odds ratio; CI, confidence interval
\*replicated and known HCM loci

**Table 2.**
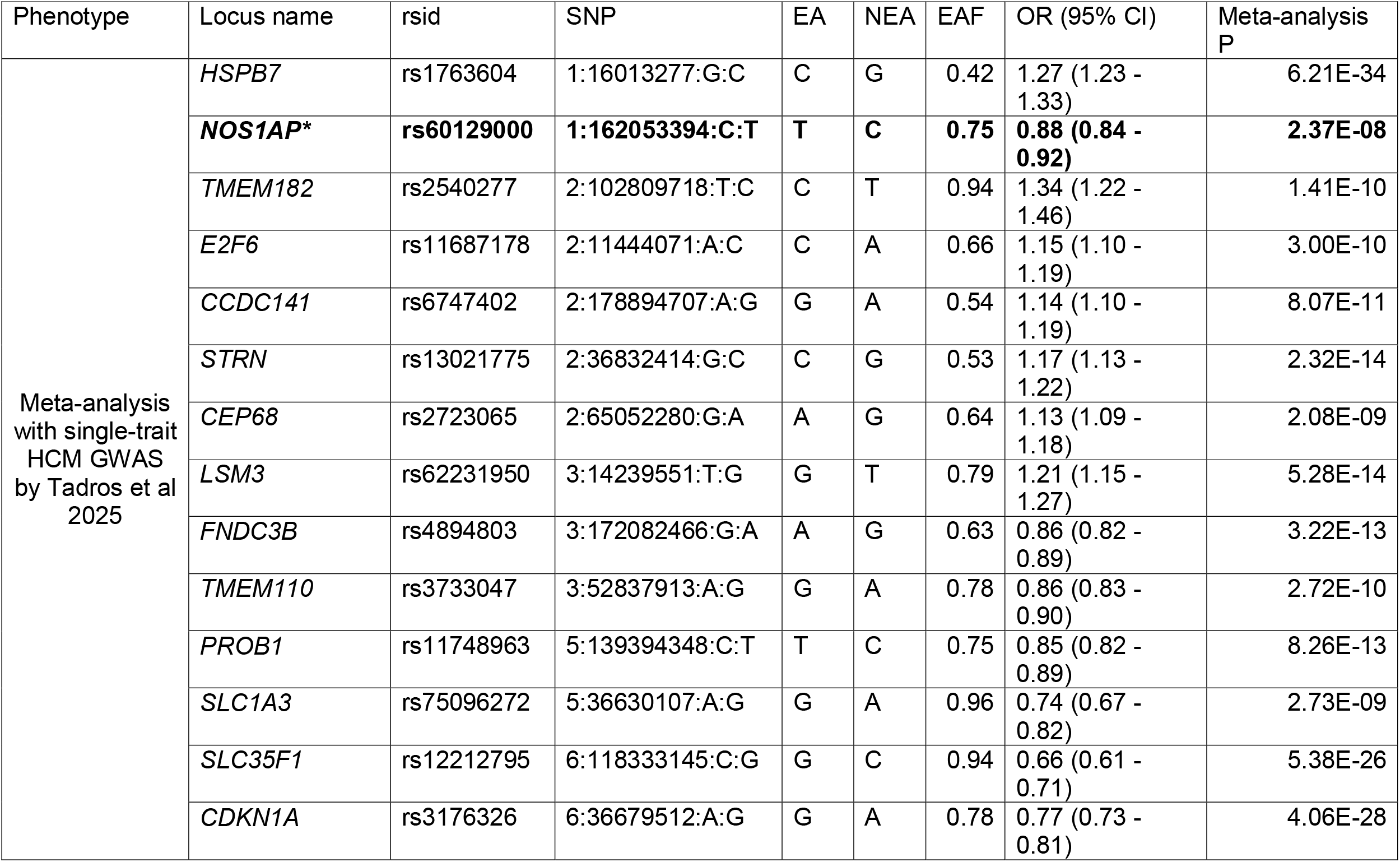
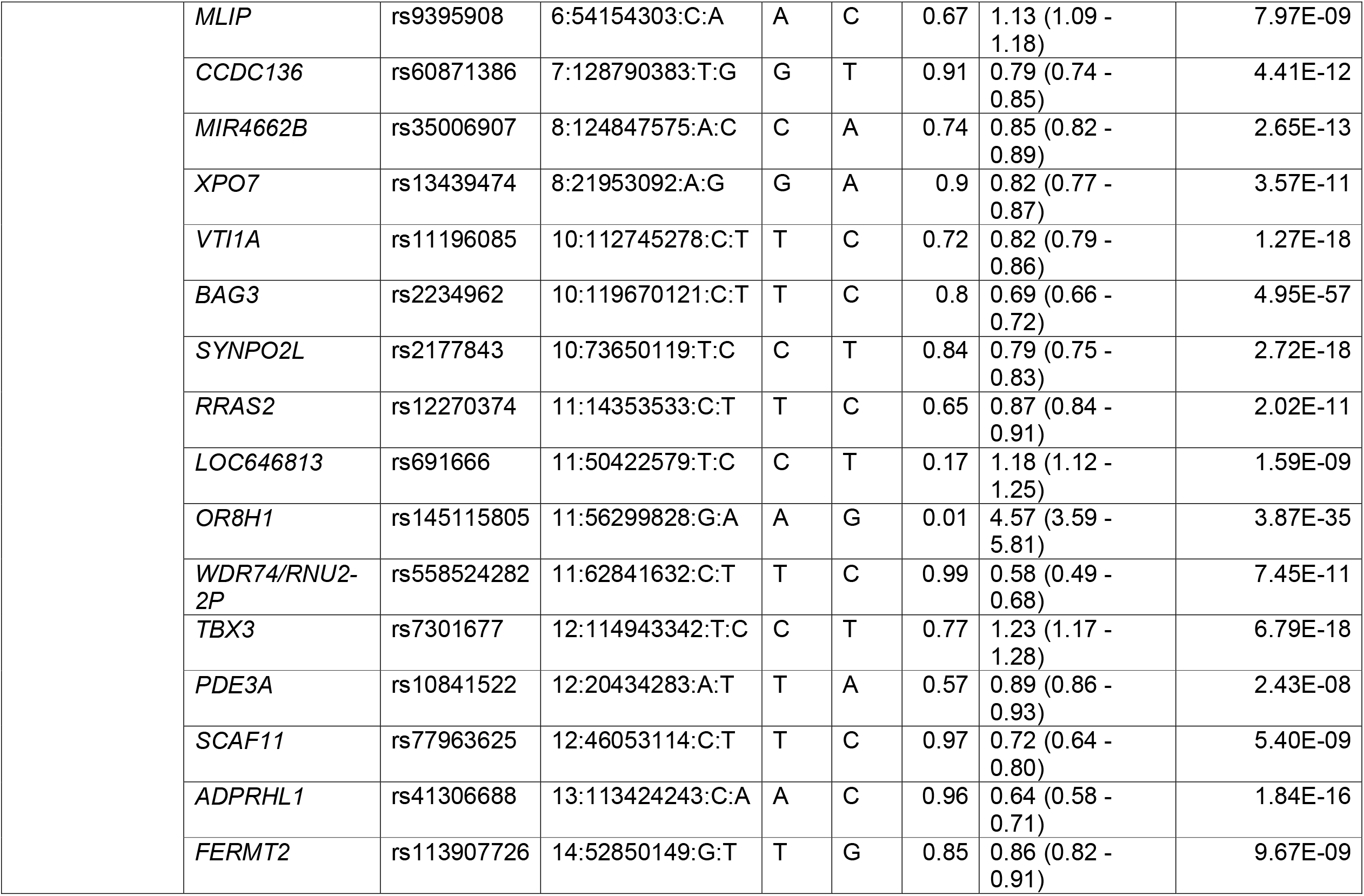
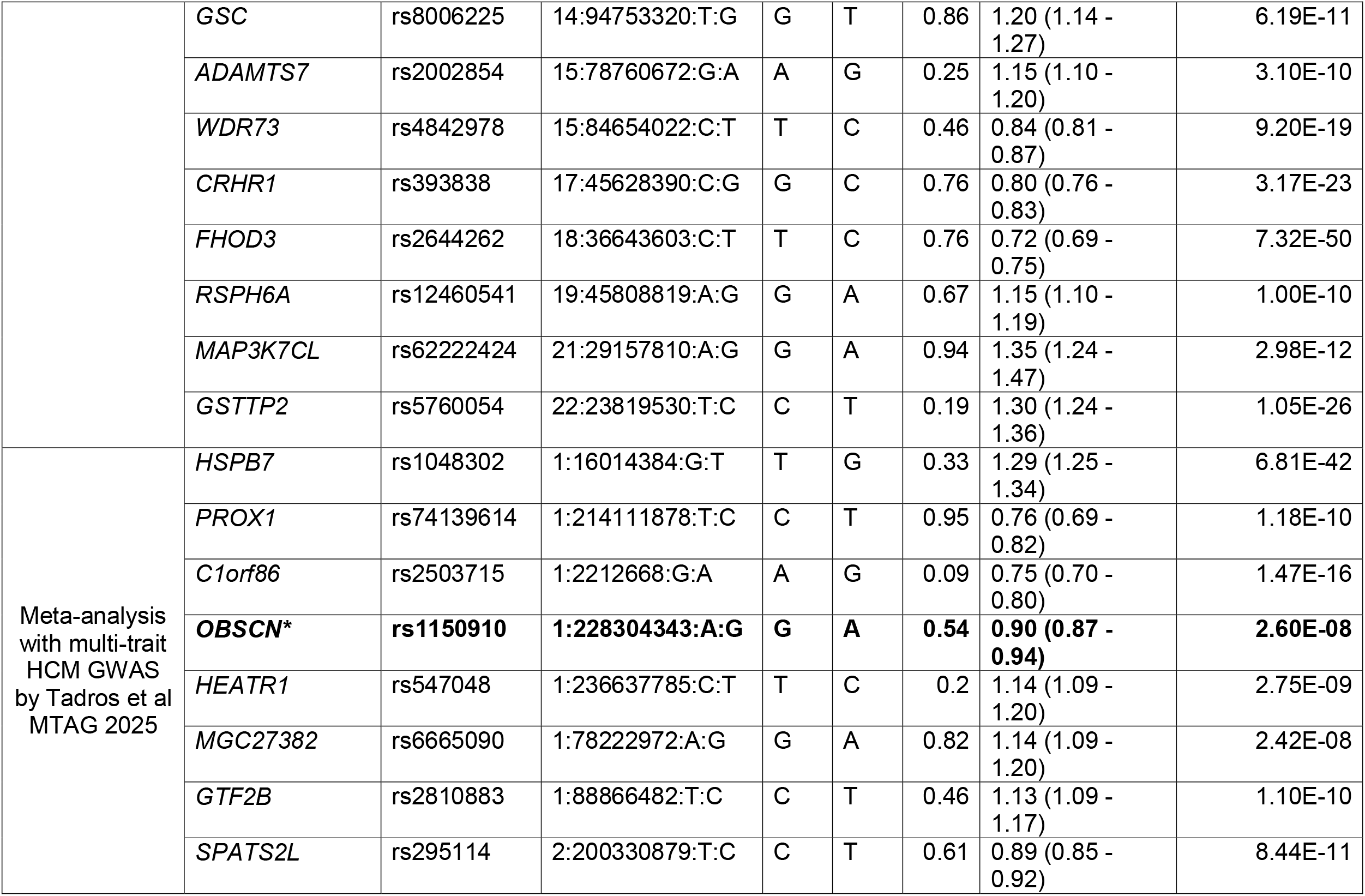
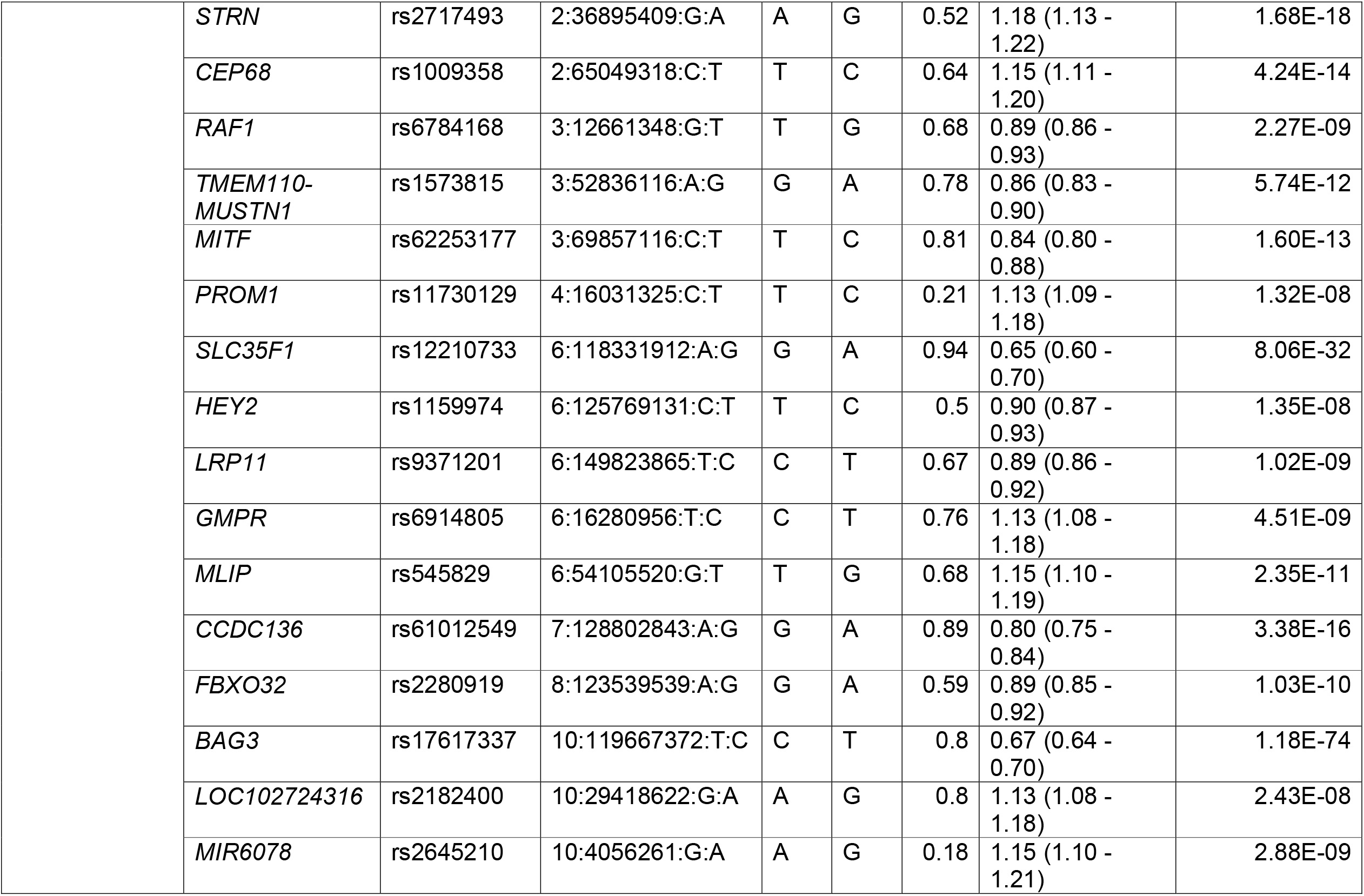
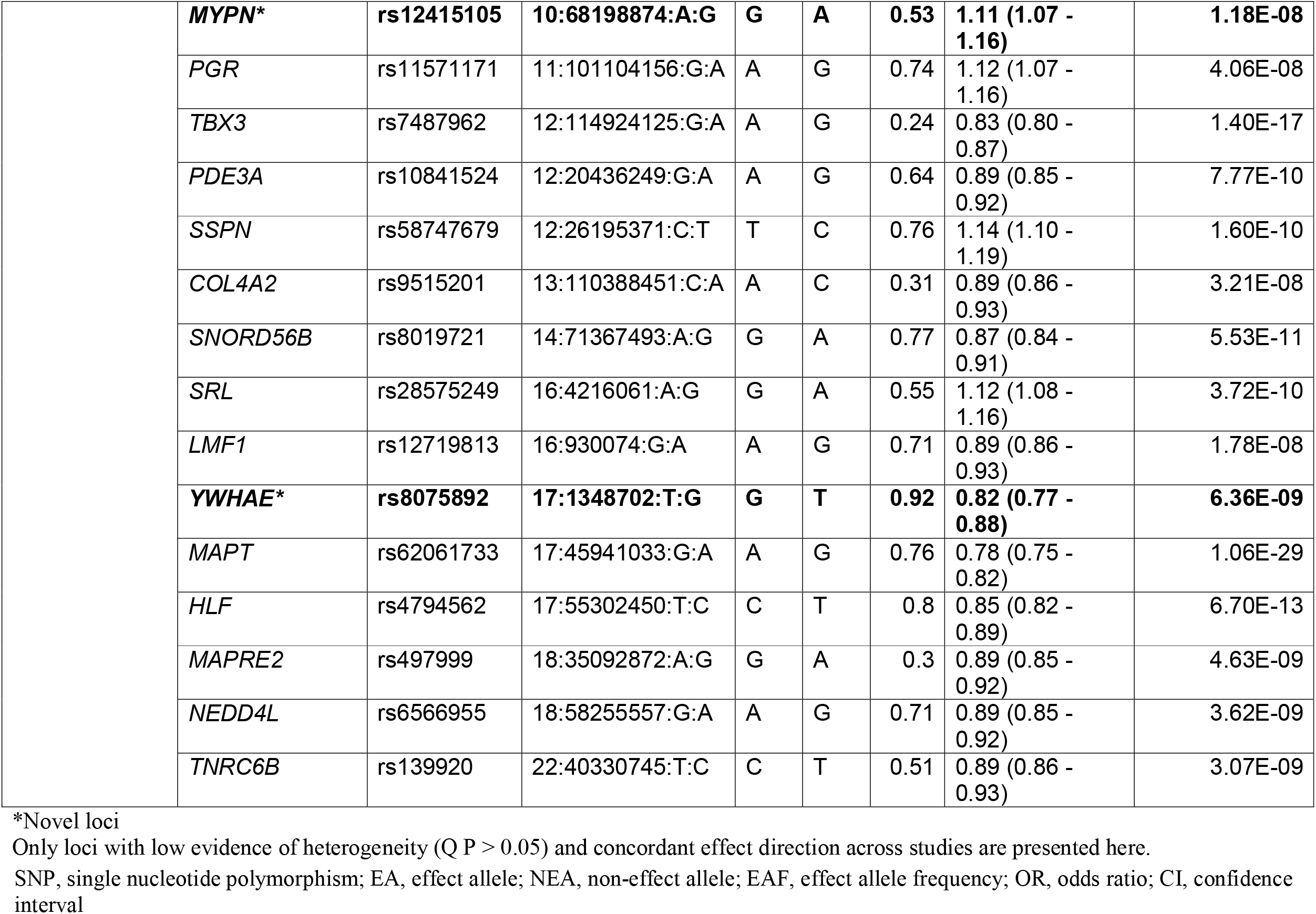
HCM GWAS meta-analysis loci.

**Figure 2.**
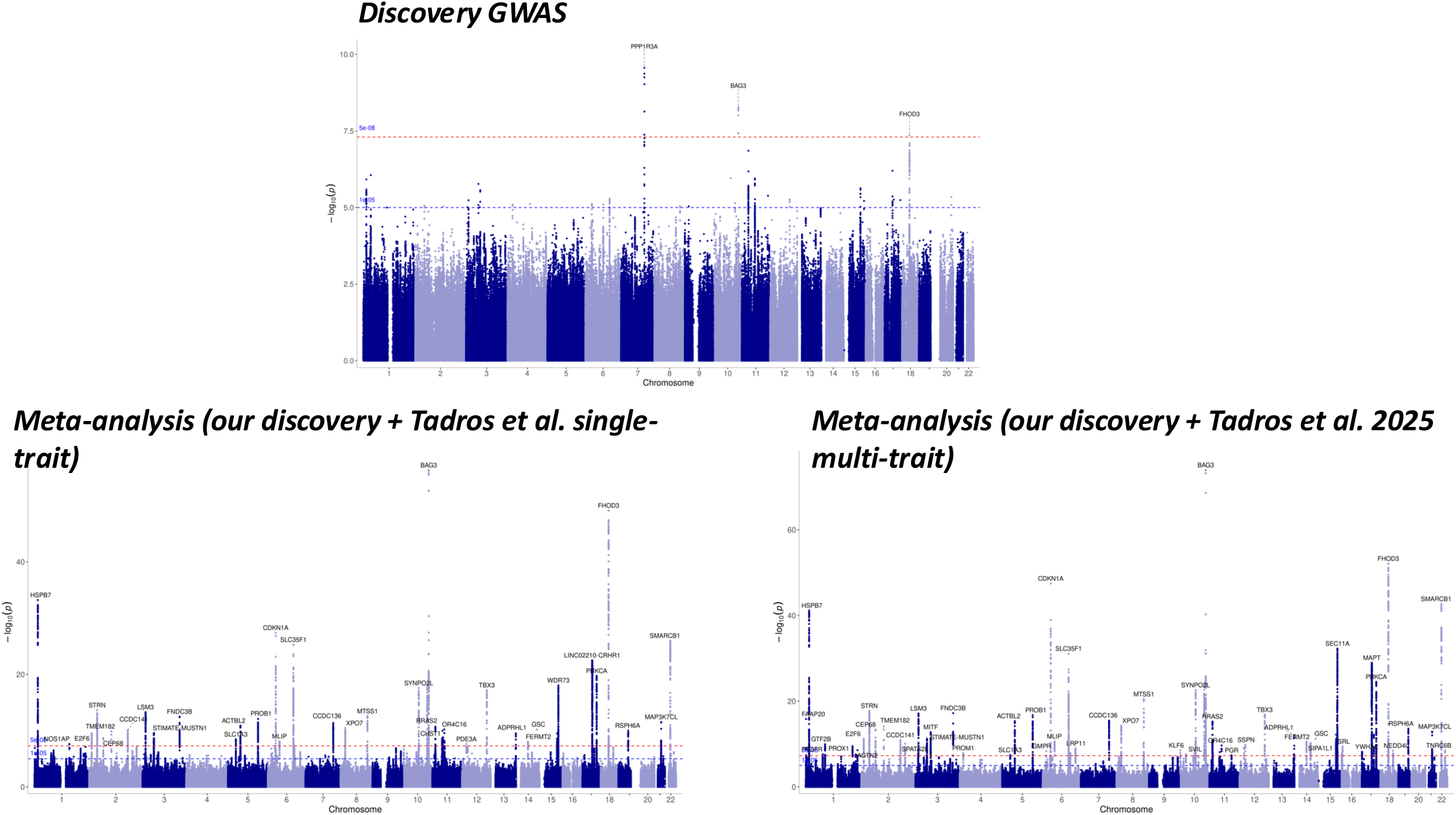
Manhattan plot of HCM loci. Each point represents a genetic variant. The red line indicates the genome-wide significant threshold at P < 5x10^-8^ and the blue line indicates the suggestive significance at P < 1x10^-5^ statistics. . P values are two-sided based on the Chi-square test

### Meta-analysis with recent Tadros et al dataset

We meta-analysed our summary statistics with single and multi-trait GWAS summary statistics from Tadros et al. (in 5,900 HCM cases and 68,359 controls). Across two meta-analyses with Tadros et al. summary statistics, in total 70 unique loci were identified (meta-analysis P < 5x10^-8^ and non-significant heterogeneity, Q P > 0.05) (**Table 2** and **Supplementary Table 3**), two of which were also discovery loci (BAG3 and FHOD3). The third discovery locus, *PPP1R3A*, reached nominal significance in the single-trait meta-analysis (OR = 0.71 [95% CI = 0.63-0.79], P^discovery^ = 2.75x10^-10^; P _meta_ = 8.0x10^-5^), with significant heterogeneity (Q P < 0.05). Thirty-eight loci out of the 70 loci were discovered by meta-analysing with the single-trait meta-analysis by Tadros et al. and 32 were additionally discovered by meta-analysing with their MTAG study. Four loci were novel HCM GWAS loci (*NOS1AP* from the single-trait meta-analysis, and *OBSCN, MYPN*, and *YWHAE* from the multi-trait meta-analysis). In these GWAS meta-analyses, there was limited genomic inflation, much of which was resolved as polygenic signal (λ = 1.155 and λ =1.149, and LD score regression intercepts 1.03 and 0.97, for single and multi-trait meta-analysis respectively). Q-Q plots for meta-analysis are shown in **Supplementary Figure 4**.

### Look-up of HCM GWAS lead variants in our discovery GWAS

We assessed if previously reported HCM loci had evidence of support in our HCM discovery analysis. Among 13 lead variants in loci identified by in Harper et al ^7^, 7 were replicated with concordant effect direction at Bonferroni adjusted P < 4.16 x 10^-3^; 4 were P < 0.05 with concordant effect direction. One lead variant (rs118060942, in *FHOD3*) was not found in our GWAS and one locus (*PRKCA*) had no evidence of support in our discovery analysis (**Supplementary Table 4**). Eight out of 16 loci described by Tadros et al (2021) ^8^ were replicated at Bonferroni adjusted P < 3.1 x 10^-3^ in our discovery GWAS. Among the eight lead variants that did not replicate, four (*SMARCB1, VTI1A, ZNF592*, and *CCDC136*) had nominal P values of <0.05 with concordant effect direction in our GWAS; however, the remaining four loci (*PKD1, PRKCA, FNDC3B*, and *ACTBL2*) had no evidence of support (P > 0.05) (**Supplementary Table 5**).

We also investigated the significance and effect direction of the 74 loci from Tadros et al. 2025 HCM GWAS in our discovery GWAS (**Supplementary Table 6**). Ten out of 74 loci were concordantly replicated at Bonferroni adjusted P < 6.7 x 10^-3^ (*HSPB7, CEP68, CDKN1A, SLC35F1/PLN, MTSS1, MYOZ1/SYNPO2L, BAG3, ADAMTS7/MORF4L1, FHOD3/TPGS2*, and *CCT8*). Twenty-five had nominal significance (P < 0.05) and concordant effect direction.

### Functional annotation of GWAS variants

We performed fine-mapping and annotation of variants in credible sets or close proxies to lead variants (LD r^2^ ≥0.8) for all loci from the meta-analyses (with P < 5x10^-8^ and Q P-value > 0.05). Annotation found 4,255 candidate variants – 0.67% were exonic, 46.02% intronic, 5.33% intergenic, and the remaining were non-coding RNA, untranslated, upstream and downstream variants (**Supplementary Table 7**). Ten loci were predicted as having damaging consequences via CADD and RegulomeDB (Methods) (*MYPN, SYNPO2L, FERMT2, SEC11A, CRHR1, MAPT, FNDC3B, TMEM110, PROB1*, and *CCDC136*). We also interrogated regulatory variants that might affect gene expression levels of their target genes in heart and arterial tissue by interrogating publicly available eQTL data-sets from the GTEx database (v8). Colocalisation in cardiovascular tissues indicated 27 putative causal variants (H4 > 0.8) in 19 loci with 17 genes in close proximity (**Supplementary Table 7**). Notably, rs11748963 (*PROB1*) colocalised with gene expression in the aorta, the atrial appendage and the left ventricle (PP4 > 0.8) and was annotated as a potentially damaging missense variant with a likely regulatory role (CADD > 20, RegulomeDB ≤ 1f and RegulomeDB probability > 0.75). 12 other variants also colocalised with more than one cardiovascular tissue, including rs12143842, in the novel *NOS1AP* locus (β = 0.1232, SE = 0.0222, OR = 1.13 (95% CI = 1.08– 1.18), P = 2.80x10^-8^), which colocalised with expression in the atrial appendage and left ventricle. rs12143842 also significantly upregulates *NOS1AP* eQTL expression in the atrial appendage and the left ventricle in GTEx.

In addition, genetic variants may have a causal effect through regulatory chromatin interactions. We identified variants regulatory potential (RegulomeDB score ≤ 5 and RegulomeDB probability > 0.75) pointing to rs12899940, rs12907511 and rs11633519 (all annotated to *CHRNB4*) in the *ADAMTS7* locus whose promoter regions also form significant interactions in tissues from the aorta and left ventricle using publicly available Hi-C data (**Supplementary Table 7**). Amongst the four novel loci across the meta-analysis, seven variants also have potential regulatory roles (RegulomeDB score ≤ 5 and RegulomeDB probability > 0.75) within the *OBSCN* (rs287610 and rs6656520), *MYPN* (rs2101483, rs7922909, rs10823155, rs10998009) and *YWHAE* (rs73292845) loci.

### Genetic overlap with other traits

Interrogation of the GWAS catalog (**Supplementary Table 8**) using the meta-analysis loci variants and their proxies (LD r^2^>=0.8) revealed associations with 438 traits, 38 of which were cardiovascular traits, in 35 loci. Variants in 34 loci had known associations with HCM in previous studies, with only the *LMF1* locus associating only with BMI. Additionally, 28 loci had variants that were also associated with the imaging measurements of left or right ventricular structure or function.

From the novel loci in our meta-analysis, *NOS1AP* and *YWHAE* had associations with cardiovascular traits. *NOS1AP* had associations with 11 cardiovascular traits (all electrocardiographic traits except one association with “Magnetic Resonance Imaging of the Heart” [rs12143842]). *YWHAE* had associations with heart failure (rs117510670, ^28^), atrial fibrillation (rs74776107, PMID: 40050429), and left ventricular mass to end-diastolic volume ratio (rs12452627, ^29^)^3030^(30)(34)(34)(34)(34)(34)(34)(35).

### Gene prioritisation and pathway enrichment

We analysed genes within 10kb of all our loci via the polygenic priority score (PoPS), which leverages gene-level association data from GWAS summary statistics to assess polygenic enrichment across gene features, including cell-type-specific expression, biological pathways, and protein-protein interactions. Among 318 genes harboured by our meta-analysis loci within a 10kb window, we utilised a custom gene prioritisation scoring system, integrating various bioinformatic resources to enhance the evidence supporting gene selection (**Supplementary Table 9**). The prioritisation framework integrated: (1) PoPS, (2) annotations from OMIM or ClinGen related to cardiovascular disease, or cardiovascular phenotypes in mouse models, and (3) evidence of enrichment in GTEx cardiovascular tissues or Hi-C data suggesting cardiac chromatin interactions. Each of the three criteria was given equal weight. Genes supported by all three evidence sources received a score of 3, whereas those supported by only one were assigned a score of 1. Ten genes had a score of 3 (*BAG3, FHOD3, NOS1AP, CEP68, FNDC3B, CDKN1A, FLNC, ADAMTSL3, GTF2B*, and *RAF1*), 38 were given a score of 2, and 110 were given the lowest score of 1. The genes prioritised at score 3 included established Mendelian cardiovascular disease genes (*FHOD3, FLNC*, and *BAG3*). The four novel loci across the meta-analysis had 23 genes nearby in total, *NOS1AP* was prioritised at the highest score of three, two novel loci had genes assigned a prioritisation score of two (with one being a known cardiomyopathy gene, *MYPN*, and *C1orf35* in the *OBSCN* locus), and seven genes in these loci were assigned the score of 1.

We also investigated pathway and mouse model enrichment of all 158 prioritised genes with score ≥ 1 using DEPICT and IMPC (**Supplementary Tables 10 and 11**). DEPICT identified prioritised genes significantly enriched for cardiovascular phenotypes (e.g. ‘increased heart weight’), GO terms (e.g., ‘heart development’) and KEGG pathways for cardiomyopathies (**Supplementary Table 10**). DEPICT analysis also identified significantly enriched pathways such as the TEAD1 (transcriptional enhancer factor domain family member 1) PPI subnetwork (P=1.89x10^-7^) which has been shown to promote cardiac remodelling ^31^. The 158 prioritised genes were also enriched for 75 knockout mouse model phenotypes (20 of which were cardiovascular-related phenotypes), with the most significant being “enlarged heart” (P=1.92x10^-10^), “abnormal retina blood vessel morphology” (P=3.26x10^-07^) followed by “abnormal heart morphology” (P=5.46x10^-07^) (**Supplementary Table 11**). The four novel loci had genes enriched for KEGG pathways such as “Hippo signaling pathway” (including *YWHAE, WNT3A*, and *WNT9A*, adjusted p-value =0.039) (**Supplementary Table 12**) which has been shown to impact mitochondrial dysfunction in DCM (https://pmc.ncbi.nlm.nih.gov/articles/PMC8419046/) ^32^. Other enriched pathways were primarily involved in oncogenic or general cellular signalling processes.

### Druggability Analysis

The druggability of annotated genes was assessed using Open Targets drug annotations via ChEMBL. 20 genes were drug targets (**Supplementary Table 9**), 14 of which were targets of cardiovascular drugs (including antihypertensives, antiarrhythmics, anticoagulants, and heart failure medications). The other six genes targeted only non-cardiovascular disease drugs, (*CHRNA5, CRHR1, COL4A2, FOLH1, MAPT*, and *ALAS1*) - targeting 26 drugs in total, including conditions such as mental illness and cancer. Out of these six genes with only non-cardiovascular drug interactions, *MAPT* was also significantly enriched for “polycystic kidney” (adjusted P=6.41x10^-5^) and “persistence of hyaloid vascular system” (adjusted P=8.06x10^-5^) in mouse model phenotypes.

### Polygenic risk score and risk of HCM and imaging phenotypes in UK Biobank population

We utilised the independent lead variants from our meta-analysis loci to construct a PRS and tested whether it predicted HCM diagnosis in the UKBB population (Figure 3) (**Supplementary Table 13**). This investigation found a HR of 1.88 (95% CI 1.72 – 2.06, P < 5.54x10^-45^ per SD increase in the continuous model) (**Supplementary Table 14**). Being in the top 5% of the PRS increased the risk of HCM three-fold compared to the bottom 95%, with an HR of 3.19 (95% CI 2.46 – 4.14, P = 2.48x10^-18^). This analysis was controlled for the presence of hypertension. Across 56,945 UKBB participants with imaging data and without a known HCM diagnosis, higher HCM PRS was significantly associated with higher LVEF (β = 0.71% per SD, 95% CI 0.66-0.76, P = 4.36x10^-174^) and with greater maximal wall thickness (β = 0.13 mm per SD, 95% CI 0.12-0.14 P = 8.89x10^-101^). (**Supplementary Table 15**) There was no evidence of association with LA size.

**Figure 3.**
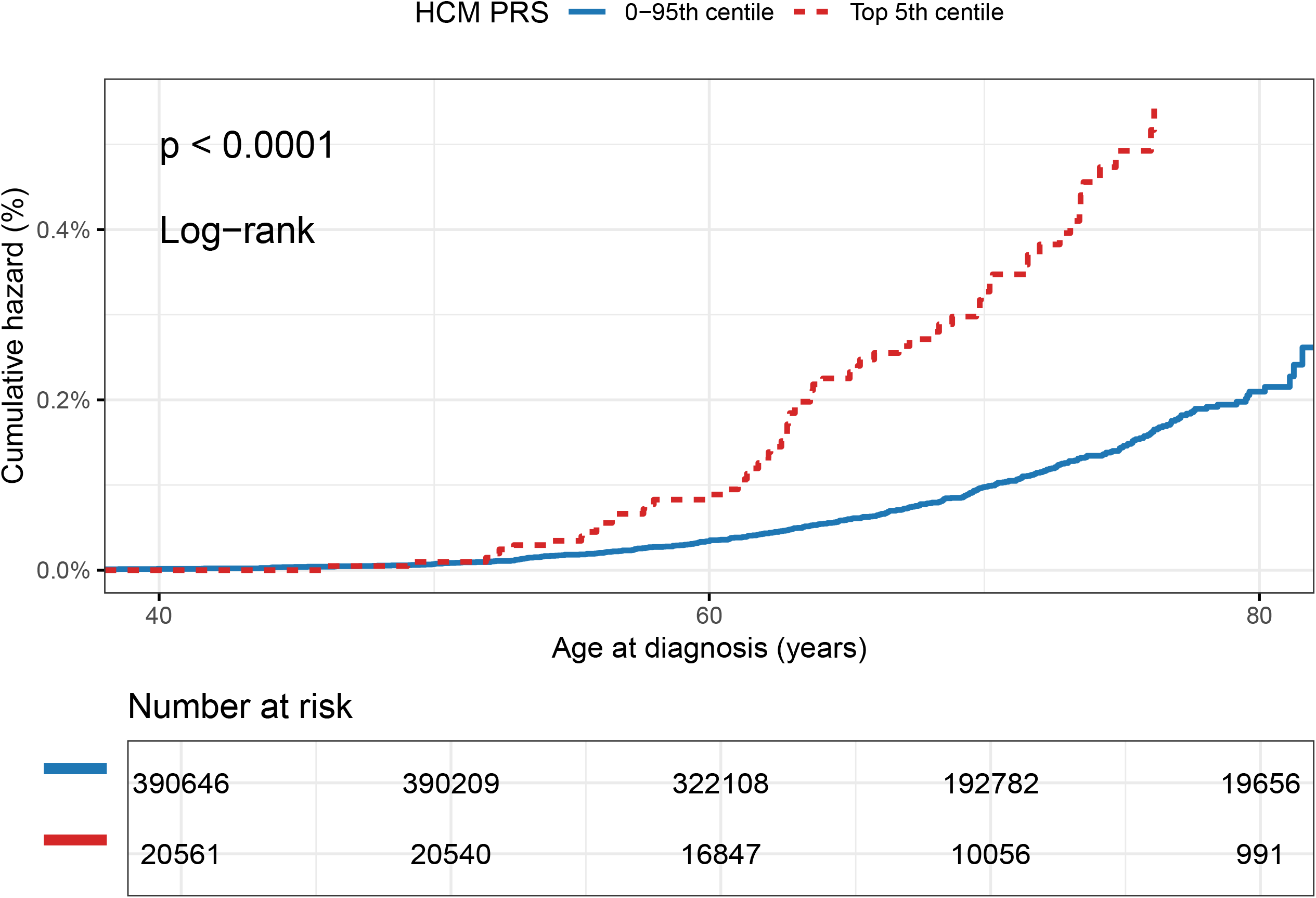
Incidence of HCM in the UKBB cohort according to PRS quintile (top 5% vs bottom 95%). Graph shows the incidence of HCM in the bottom 95% (blue) compared to the top 5% PRS scores (red). The two curves diverged significantly according to the log rank test.

### Association of PRS with intermediate phenotypes and outcomes in HCM cases, stratified according to the presence or absence of a sarcomere causal variant

There was a significant positive association between LVEF and HCM PRS in the discovery HCM sarcomere-positive cases (effect size 2.4%, 95% CI: 1.28 – 4.51%, P = 0.006) (**Supplementary Table 16**). For clinical outcomes, no significant associations were observed per SD increase in PRS; however, when comparing individuals in the top 20% versus the bottom 80% of the PRS distribution, a higher PRS was associated with an increased risk of sudden cardiac death or equivalent among sarcomere-negative HCM cases (HR 2.72, 95% CI: 1.03–7.17; P = 0.043) (**Supplementary Table 17**).

## DISCUSSION

We have conducted a case-control GWAS comprising more than 2,000 HCM samples not previously studied for a polygenic contribution to HCM and discovered three loci, of which one was novel. Meta-analysis of our data with a recently published GWAS of single and multi-trait HCM analyses identified a total of 70 unique genome-wide significant loci including four novel loci. Our HCM polygenic risk score was associated with the risk of HCM and its imaging features in UKBB. A higher PRS was associated with an increased risk of sudden cardiac death or equivalent among sarcomere-negative HCM cases.

### Novel loci and genes associated with HCM

Stimulated by the incomplete yield of genetic testing in HCM, recent work has tested the hypothesis that common variation is the cause of some genotype-elusive cases. Three publications have indeed described a number of loci associated with HCM and reported heritability for common variation ranging from 0.12-0.29 ^8^, 0.14-0.36 ^7^ and 0.16-0.29 ^11^ with the highest values in sarcomere-negative cases. However, there is still substantial missing heritability to be explored.

Our discovery GWAS reported three loci including a novel HCM locus (*PPP1R3A*). The meta-analyses provided some support for the novel HCM locus. It worth commenting that there has been inconsistency in the ability to validate HCM loci across the different studies. For example, we could not reproduce some of the loci described in previous GWAS, such as *PKD1, PRKCA, FNDC3B*, and *ACTBL2*.

The *PPP1R3A* gene (discovery P=2.75x10^-10^, meta-analysis P=8.05x10^-5^ and Q P < 0.05), is an interesting gene at the novel locus. The gene encodes Protein Phosphatase 1 Regulatory Subunit 3A, recently identified as a potential central regulator in a cardiac regulatory gene networks analysis of failing hearts ^33^. *In vitro* knockdown of *PPP1R3A* revealed a role in metabolic pathway regulation and an association with cardiomyocyte size and perturbed respiratory metabolism. Murine *ppp1r3a* knockouts were protected against pressure-overload heart failure ^33^. Other work has shown reduced levels of *PPP1R3A*, increased phosphorylation of phospholamban (PLN), a new HCM locus from this workand ryanodine receptor RyR2, leading to abnormal sarcoplasmic calcium release, promoting atrial fibrillation ^34^; PPP1R3A was suggested to be part of a macromolecular complex including PLN, RyR2 and SERCA2A. We report this locus and candidate gene, but note this discovery data needs to be interpreted carefully given the consistent albeit heterogeneous signal from the meta-analysis.

The meta-analysis data provided four additional novel loci with non-significant heterogeneity: *NOS1AP, OBSCN, MYPN*, and *YWHAE*. All loci have candidate genes which are biologically plausible for HCM.

Truncating and missense variants in *MYPN* (myopalladin)–a Z-disc protein–have been associated with DCM but association of rare variants with HCM has not been definitely established ^35^. The same is true for *OBSCN* (obscurin), another Z-disc associated protein with a role in sarcomere integrity. Functional work and sequencing studies have suggested an association with HCM and DCM that remains disputed ^35^, however, the association found here, alongside its suggestive significant association in Tadros et al. (P = 0.000062) warrants further investigation.

*NOS1AP* (Nitric Oxide Synthase 1 Adaptor Protein), at the *NOS1AP* locus, had a high score from our gene prioritisation scoring system, reflecting evidence from various lines of downstream bioinformatic analysis. In addition, *NOS1AP* polymorphisms have been associated with QT duration in HCM ^36^, although the specific mechanisms involved are not known.

*YWHAE* (Tyrosine 3-Monooxygenase/Tryptophan 5-Monooxygenase Activation Protein Epsilon) is a transcriptional regulator of the cardiac voltage-gated sodium channel alpha subunit 5 (Na_V_ 1.5) (encoded by *SCN5A*) ^37^ and associated with left ventricular end-diastolic mass/volume ratio in a recent GWAS ^29^.

From our druggability analysis, six previously reported HCM genes have only non-cardiovascular drug interactions: *CHRNA5, MAPT, CRHR1, COL4A2, FOLH1*, and *ALAS1*. However, for all of these genes, except for *FOLH1*, research has reported a link to the regulation of cardiovascular structure and function with notably a *CRHR1* agonist reducing left ventricular ejection fraction in mice, and *COL4A2* knockout mice having increased vascular wall thickening. However, these genes also have known roles outside of the cardiovascular system that indicate strong pleiotropic effects (**Supplementary Table 8**); for example *CHRNA5* locus is notable for its pleiotropic associations particularly with cigarettes smoked per day, as well as several cardiovascular-related traits (e.g., coronary artery disease, stroke, chronic kidney disease, and blood pressure). Therefore, its drug interactions may reflect indirect effects via smoking behaviour or pleiotropic mechanisms, rather than a direct causal role in cardiovascular pathology.

### A new polygenic risk score predicted HCM in UK Biobank

We constructed a HCM PRS using the lead variants from our meta-analysis and tested its validity by predicting HCM diagnosis in the UKBB population. A significant association was found, with an HR of 1.81 per 1SD increment in the PRS. Our associations were independent of hypertension. This is relevant as hypertensive heart disease can be misdiagnosed as HCM, particularly in older individuals. A recent publication obtained reported an odds ratio (OR) of 1.97 per 1SD increment, (95% CI⍰=⍰1.81–2.15, P⍰<⍰2⍰×⍰10^−16^), which is similar to our results ^9^. Importantly, an association was also observed between PRS and imaging phenotypes in UKBB participants without a diagnosis of HCM.

### Associations with phenotype and outcomes

A broad composite endpoint including the incidence of atrial fibrillation, arrhythmic events, heart failure events and mortality was associated with PRS above the median in a previous publication ^8^. In a more recent study, HCM PRS predicted adverse outcomes including death and the risk of developing HCM in relatives ^9^. In our study, sarcomere-negative individuals in the top 20% of our PRS had association with increased risk of SCD or equivalent. We also detected a marginal association with LVEF in the HCM cohort.

### Limitations and implications for future work

The absence of more significant associations with outcomes could be due to limitations in statistical power with the small HCM sample. While we identified novel loci, significant heterogeneity across cohorts used for the meta-analysis reduced the reliability of some signals. Some previously published loci were not able to be validated. These inconsistencies likely reflect differences between cohorts in case ascertainment (including phenotype definition) and allele frequency spectra, underscoring the need for cautious interpretation of individual associations in any GWAS study. As the data in this study derives from adults, the association of PRS with incidence of HCM might not be generalisable to children and young adults. Although our discovery case-control GWAS did not include the UKBB cohort, it contributed both to the published GWAS summary statistics used in our meta-analyses and to the evaluation of our HCM PRS, raising the possibility of partial sample overlap. Replication in fully independent cohorts is needed to confirm the predictive capability of this specific PRS, alhough results are aligned with data reported in previous publications. Our study cohort was of European ancestry, it will be important that future work pools multi-centre cohorts, including non-European ancestries, and look to extend follow-up to improve power.

## Conclusions

We report novel loci associated with HCM and demonstrate the association between HCM PRS and an increased risk of HCM, imaging phenotypes in UKBB and outcomes in HCM patients.

## Data Availability

All data produced in the present study are available upon reasonable request to the authors

## Acknowledgments

EXCEED is supported by the University of Leicester, the NIHR Leicester Respiratory Biomedical Research Centre, by the Wellcome Trust [WT225221/Z/22/Z], and by Cohort Access fees from studies funded by the Medical Research Council (MRC), BBRSC, NIHR, the UK Space Agency, and GSK. The views expressed are those of the author(s) and not necessarily those of the NIHR or the Department of Health and Social Care. It was previously supported by MRC grant G0902313, by the Wellcome Trust [WT202849/Z/16/Z], and by BREATHE - The Health Data Research Hub for Respiratory Health [UKR_PC_19004] in partnership with SAIL Databank. Our analyses of EXCEED used the ALICE High Performance Computing Facilities at the University of Leicester. The EXCEED study gratefully acknowledges the support of all participants and staff who have contributed to the study. This work was partially supported by the National Institute for Health Research (NIHR) Leicester Biomedical Research Centre; the views expressed are those of the author(s) and not necessarily those of the National Health Service, the NIHR, or the Department of Health and Social Care

## REFERENCES

1. Authors/Task Force m, Elliott PM, Anastasakis A, Borger MA, Borggrefe M, Cecchi F, Charron P, Hagege AA, Lafont A, Limongelli G, et al. 2014 ESC Guidelines on diagnosis and management of hypertrophic cardiomyopathy: The Task Force for the Diagnosis and Management of Hypertrophic Cardiomyopathy of the European Society of Cardiology (ESC). Eur Heart J. 2014;35:2733–2779. doi: 10.1093/eurheartj/ehu284

2. Christian S, Cirino A, Hansen B, Harris S, Murad AM, Natoli JL, Malinowski J, Kelly MA. Diagnostic validity and clinical utility of genetic testing for hypertrophic cardiomyopathy: a systematic review and meta-analysis. Open Heart. 2022;9. doi: 10.1136/openhrt-2021-001815

3. Lopes LR, Garcia-Hernandez S, Lorenzini M, Futema M, Chumakova O, Zateyshchikov D, Isidoro-Garcia M, Villacorta E, Escobar-Lopez L, Garcia-Pavia P, et al. Alpha-protein kinase 3 (ALPK3) truncating variants are a cause of autosomal dominant hypertrophic cardiomyopathy. Eur Heart J. 2021;42:3063–3073. doi: 10.1093/eurheartj/ehab424

4. Ochoa JP, Sabater-Molina M, Garcia-Pinilla JM, Mogensen J, Restrepo-Cordoba A, Palomino-Doza J, Villacorta E, Martinez-Moreno M, Ramos-Maqueda J, Zorio E, et al. Formin Homology 2 Domain Containing 3 (FHOD3) Is a Genetic Basis for Hypertrophic Cardiomyopathy. J Am Coll Cardiol. 2018;72:2457–2467. doi: 10.1016/j.jacc.2018.10.001

5. Ingles J, Burns C, Bagnall RD, Lam L, Yeates L, Sarina T, Puranik R, Briffa T, Atherton JJ, Driscoll T, et al. Nonfamilial Hypertrophic Cardiomyopathy: Prevalence, Natural History, and Clinical Implications. Circ Cardiovasc Genet. 2017;10. doi: 10.1161/CIRCGENETICS.116.001620

6. Watkins H, Ashrafian H, Redwood C. Inherited cardiomyopathies. N Engl J Med. 2011;364:1643–1656. doi: 10.1056/NEJMra0902923

7. Harper AR, Goel A, Grace C, Thomson KL, Petersen SE, Xu X, Waring A, Ormondroyd E, Kramer CM, Ho CY, et al. Common genetic variants and modifiable risk factors underpin hypertrophic cardiomyopathy susceptibility and expressivity. Nat Genet. 2021;53:135–142. doi: 10.1038/s41588-020-00764-0

8. Tadros R, Francis C, Xu X, Vermeer AMC, Harper AR, Huurman R, Kelu Bisabu K, Walsh R, Hoorntje ET, Te Rijdt WP, et al. Shared genetic pathways contribute to risk of hypertrophic and dilated cardiomyopathies with opposite directions of effect. Nat Genet. 2021;53:128–134. doi: 10.1038/s41588-020-00762-2

9. Zheng SL, Jurgens SJ, McGurk KA, Xu X, Grace C, Theotokis PI, Buchan RJ, Francis C, de Marvao A, Curran L, et al. Evaluation of polygenic scores for hypertrophic cardiomyopathy in the general population and across clinical settings. Nat Genet. 2025;57:563–571. doi: 10.1038/s41588-025-02094-5

10. Biddinger KJ, Jurgens SJ, Maamari D, Gaziano L, Choi SH, Morrill VN, Halford JL, Khera AV, Lubitz SA, Ellinor PT, et al. Rare and Common Genetic Variation Underlying the Risk of Hypertrophic Cardiomyopathy in a National Biobank. JAMA Cardiol. 2022;7:715–722. doi: 10.1001/jamacardio.2022.1061

11. Tadros R, Zheng SL, Grace C, Jorda P, Francis C, West DM, Jurgens SJ, Thomson KL, Harper AR, Ormondroyd E, et al. Large-scale genome-wide association analyses identify novel genetic loci and mechanisms in hypertrophic cardiomyopathy. Nat Genet. 2025;57:530–538. doi: 10.1038/s41588-025-02087-4

12. O’Mahony C, Jichi F, Pavlou M, Monserrat L, Anastasakis A, Rapezzi C, Biagini E, Gimeno JR, Limongelli G, McKenna WJ, et al. A novel clinical risk prediction model for sudden cardiac death in hypertrophic cardiomyopathy (HCM risk-SCD). Eur Heart J. 2014;35:2010–2020. doi: 10.1093/eurheartj/eht439

13. Lopes LR, Syrris P, Guttmann OP, O’Mahony C, Tang HC, Dalageorgou C, Jenkins S, Hubank M, Monserrat L, McKenna WJ, et al. Novel genotype-phenotype associations demonstrated by high-throughput sequencing in patients with hypertrophic cardiomyopathy. Heart. 2015;101:294–301. doi: 10.1136/heartjnl-2014-306387

14. Richards S, Aziz N, Bale S, Bick D, Das S, Gastier-Foster J, Grody WW, Hegde M, Lyon E, Spector E, et al. Standards and guidelines for the interpretation of sequence variants: a joint consensus recommendation of the American College of Medical Genetics and Genomics and the Association for Molecular Pathology. Genet Med. 2015;17:405–424. doi: 10.1038/gim.2015.30

15. Pain O, Glanville KP, Hagenaars SP, Selzam S, Furtjes AE, Gaspar HA, Coleman JRI, Rimfeld K, Breen G, Plomin R, et al. Evaluation of polygenic prediction methodology within a reference-standardized framework. PLoS Genet. 2021;17:e1009021. doi: 10.1371/journal.pgen.1009021

16. Genomes Project C, Auton A, Brooks LD, Durbin RM, Garrison EP, Kang HM, Korbel JO, Marchini JL, McCarthy S, McVean GA, et al. A global reference for human genetic variation. Nature. 2015;526:68–74. doi: 10.1038/nature15393

17. John C, Reeve NF, Free RC, Williams AT, Ntalla I, Farmaki AE, Bethea J, Barton LM, Shrine N, Batini C, et al. Cohort profile: Extended Cohort for E-health, Environment and DNA (EXCEED). Int J Epidemiol. 2019;48:1734. doi: 10.1093/ije/dyz175

18. Aung N, Lopes LR, van Duijvenboden S, Harper AR, Goel A, Grace C, Ho CY, Weintraub WS, Kramer CM, Neubauer S, et al. Genome-Wide Analysis of Left Ventricular Maximum Wall Thickness in the UK Biobank Cohort Reveals a Shared Genetic Background With Hypertrophic Cardiomyopathy. Circ Genom Precis Med. 2023;16:e003716. doi: 10.1161/CIRCGEN.122.003716

19. Turley P, Walters RK, Maghzian O, Okbay A, Lee JJ, Fontana MA, Nguyen-Viet TA, Wedow R, Zacher M, Furlotte NA, et al. Multi-trait analysis of genome-wide association summary statistics using MTAG. Nat Genet. 2018;50:229–237. doi: 10.1038/s41588-017-0009-4

20. Mbatchou J, Barnard L, Backman J, Marcketta A, Kosmicki JA, Ziyatdinov A, Benner C, O’Dushlaine C, Barber M, Boutkov B, et al. Computationally efficient whole-genome regression for quantitative and binary traits. Nat Genet. 2021;53:1097–1103. doi: 10.1038/s41588-021-00870-7

21. McLaren W, Gil L, Hunt SE, Riat HS, Ritchie GR, Thormann A, Flicek P, Cunningham F. The Ensembl Variant Effect Predictor. Genome Biol. 2016;17:122. doi: 10.1186/s13059-016-0974-4

22. Jung I, Schmitt A, Diao Y, Lee AJ, Liu T, Yang D, Tan C, Eom J, Chan M, Chee S, et al. A compendium of promoter-centered long-range chromatin interactions in the human genome. Nat Genet. 2019;51:1442–1449. doi: 10.1038/s41588-019-0494-8

23. Boyle AP, Hong EL, Hariharan M, Cheng Y, Schaub MA, Kasowski M, Karczewski KJ, Park J, Hitz BC, Weng S, et al. Annotation of functional variation in personal genomes using RegulomeDB. Genome Res. 2012;22:1790–1797. doi: 22/9/1790 [pii] 10.1101/gr.137323.112

24. Consortium GT. The Genotype-Tissue Expression (GTEx) project. Nat Genet. 2013;45:580–585. doi: 10.1038/ng.2653

25. Giambartolomei C, Vukcevic D, Schadt EE, Franke L, Hingorani AD, Wallace C, Plagnol V. Bayesian test for colocalisation between pairs of genetic association studies using summary statistics. PLoS Genet. 2014;10:e1004383. doi: 10.1371/journal.pgen.1004383

26. Buniello A, MacArthur JAL, Cerezo M, Harris LW, Hayhurst J, Malangone C, McMahon A, Morales J, Mountjoy E, Sollis E, et al. The NHGRI-EBI GWAS Catalog of published genome-wide association studies, targeted arrays and summary statistics 2019. Nucleic Acids Res. 2019;47:D1005–D1012. doi: 10.1093/nar/gky1120

27. Pers TH, Karjalainen JM, Chan Y, Westra HJ, Wood AR, Yang J, Lui JC, Vedantam S, Gustafsson S, Esko T, et al. Biological interpretation of genome-wide association studies using predicted gene functions. Nature communications. 2015;6:5890. doi: 10.1038/ncomms6890

28. Levin MG, Tsao NL, Singhal P, Liu C, Vy HMT, Paranjpe I, Backman JD, Bellomo TR, Bone WP, Biddinger KJ, et al. Genome-wide association and multi-trait analyses characterize the common genetic architecture of heart failure. Nature communications. 2022;13:6914. doi: 10.1038/s41467-022-34216-6

29. Schmidt AF, Bourfiss M, Alasiri A, Puyol-Anton E, Chopade S, van Vugt M, van der Laan SW, Gross C, Clarkson C, Henry A, et al. Druggable proteins influencing cardiac structure and function: Implications for heart failure therapies and cancer cardiotoxicity. Sci Adv. 2023;9:eadd4984. doi: 10.1126/sciadv.add4984

30. Scholl UI. Genetics of Primary Aldosteronism. Hypertension. 2022;79:887–897. doi: 10.1161/HYPERTENSIONAHA.121.16498

31. Song S, Zhang X, Huang Z, Zhao Y, Lu S, Zeng L, Cai F, Wang T, Pei Z, Weng X, et al. TEA domain transcription factor 1(TEAD1) induces cardiac fibroblasts cells remodeling through BRD4/Wnt4 pathway. Signal Transduct Target Ther. 2024;9:45. doi: 10.1038/s41392-023-01732-w

32. Wu W, Ziemann M, Huynh K, She G, Pang ZD, Zhang Y, Duong T, Kiriazis H, Pu TT, Bai RY, et al. Activation of Hippo signaling pathway mediates mitochondria dysfunction and dilated cardiomyopathy in mice. Theranostics. 2021;11:8993–9008. doi: 10.7150/thno.62302

33. Cordero P, Parikh VN, Chin ET, Erbilgin A, Gloudemans MJ, Shang C, Huang Y, Chang AC, Smith KS, Dewey F, et al. Pathologic gene network rewiring implicates PPP1R3A as a central regulator in pressure overload heart failure. Nature communications. 2019;10:2760. doi: 10.1038/s41467-019-10591-5

34. Alsina KM, Hulsurkar M, Brandenburg S, Kownatzki-Danger D, Lenz C, Urlaub H, Abu-Taha I, Kamler M, Chiang DY, Lahiri SK, et al. Loss of Protein Phosphatase 1 Regulatory Subunit PPP1R3A Promotes Atrial Fibrillation. Circulation. 2019;140:681–693. doi: 10.1161/CIRCULATIONAHA.119.039642

35. Noureddine M, Gehmlich K. Structural and signaling proteins in the Z-disk and their role in cardiomyopathies. Front Physiol. 2023;14:1143858. doi: 10.3389/fphys.2023.1143858

36. Earle N, Ingles J, Bagnall RD, Gray B, Crawford J, Smith W, Shelling AN, Love DR, Semsarian C, Skinner JR. NOS1AP Polymorphisms Modify QTc Interval Duration But Not Cardiac Arrest Risk in Hypertrophic Cardiomyopathy. J Cardiovasc Electrophysiol. 2015;26:1346–1351. doi: 10.1111/jce.12827

37. Hu Y, Zhang C, Wang S, Xiong H, Xie W, Zeng Z, Cai H, Wang QK, Lu Z. 14-3-3epsilon/YWHAE regulates the transcriptional expression of cardiac sodium channel Na(V)1.5. Heart Rhythm. 2024;21:2320–2329. doi: 10.1016/j.hrthm.2024.05.015

