## Supplemental figures for "Hypertrophic cardiomyopathy: a genome wide association meta-analysis and polygenic risk score"

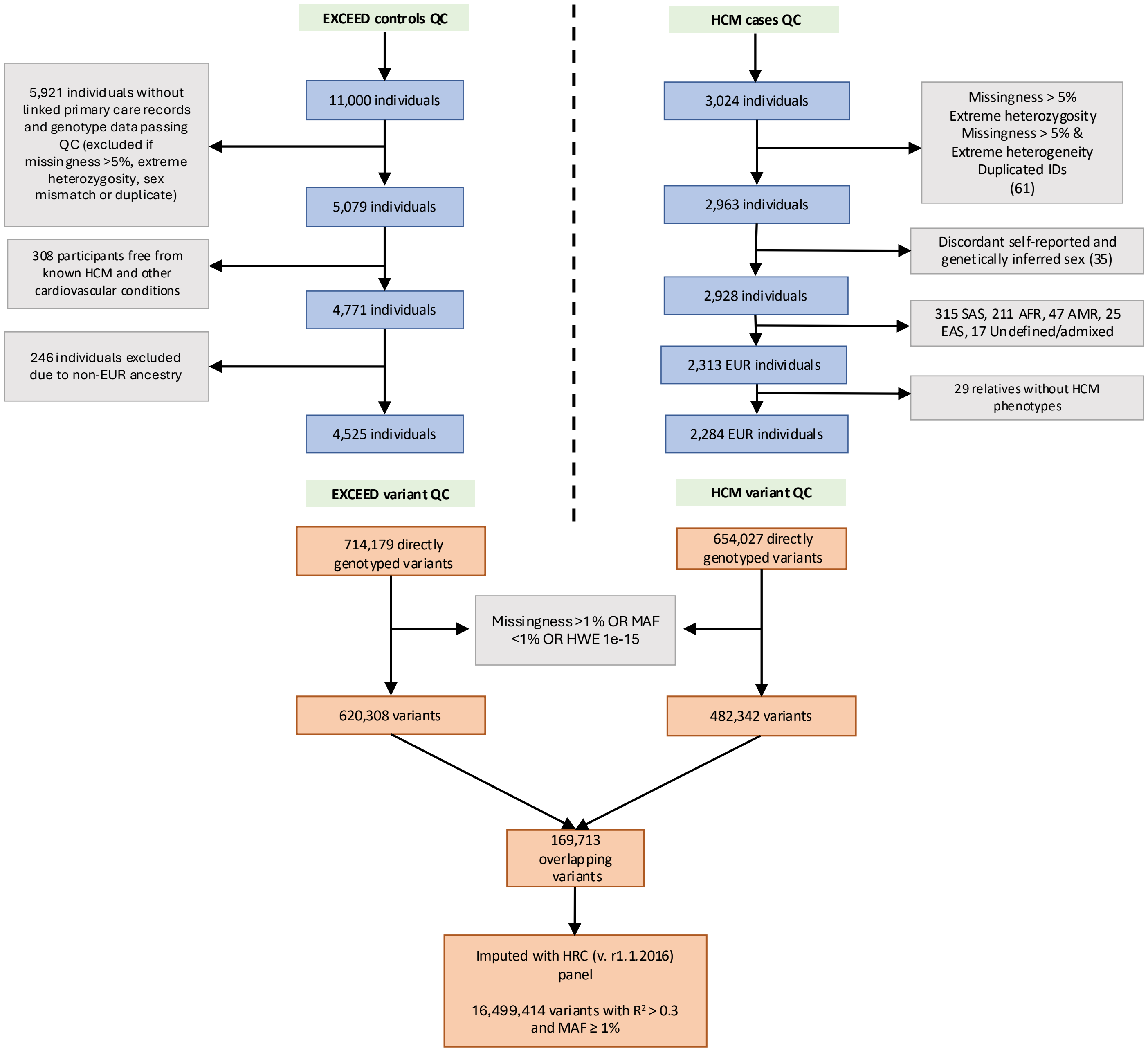


Supplementary Figure 1. Flow chart of A) Sample quality control (QC) and B) Variant QC

*HCM, hypertrophic cardiomyopathy; UKB, UK Biobank; EUR, European; SAS, South Asian; AFR, African; AMR, Admixed American; EAS, East Asian; MAF, minor allele frequency; HWE, Hardy-Weinberg equilibrium; HRC, Haplotype Reference Consortium*

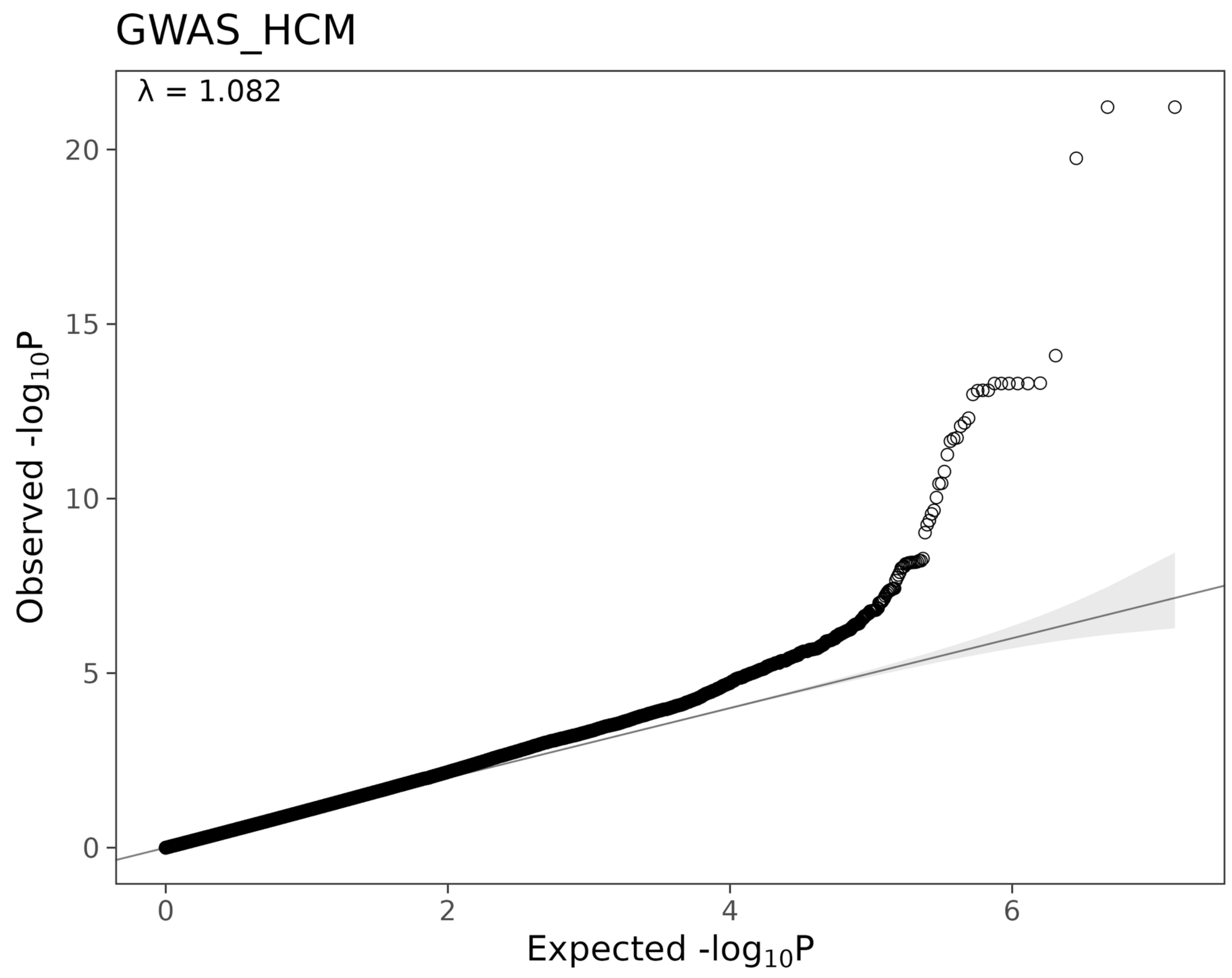

GWAS_HCM
λ = 1.082
20
15
10
5
0
Observed -log10P
0
2
4
6
Expected -log10P

### *HCM Discovery Loci*

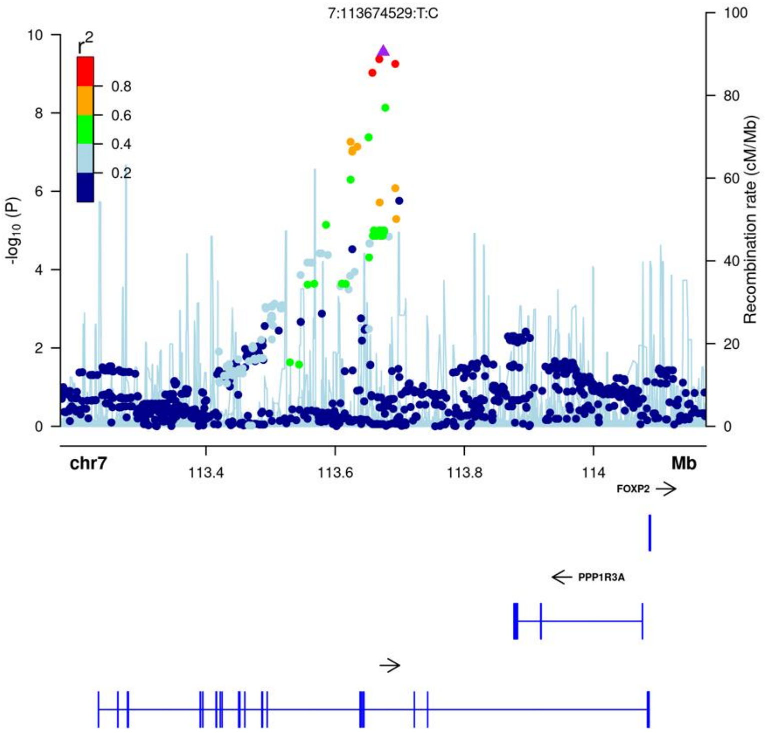


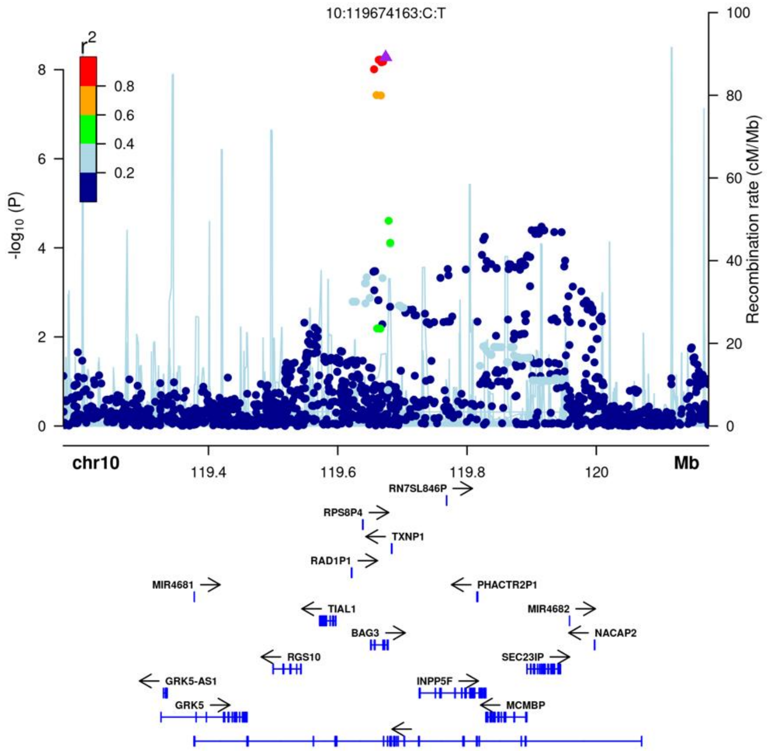


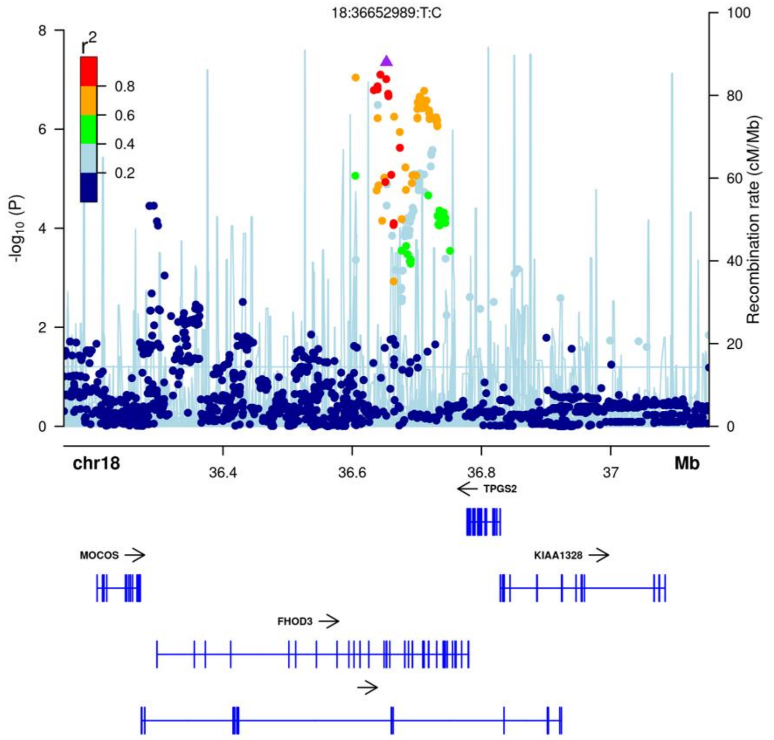

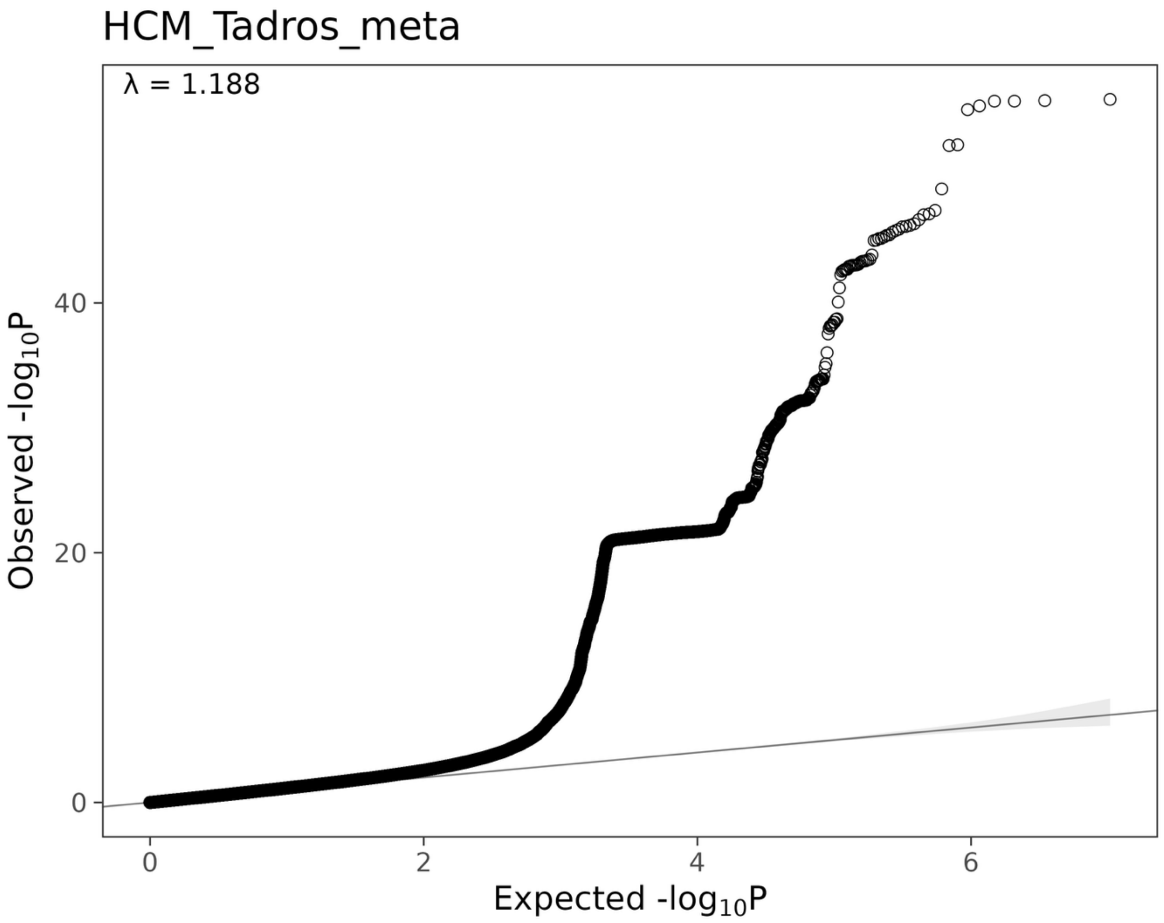
HCM_Tadros_meta
λ = 1.188
Observed -log10P
Expected -log10P
0
20
40
0
2
4
6

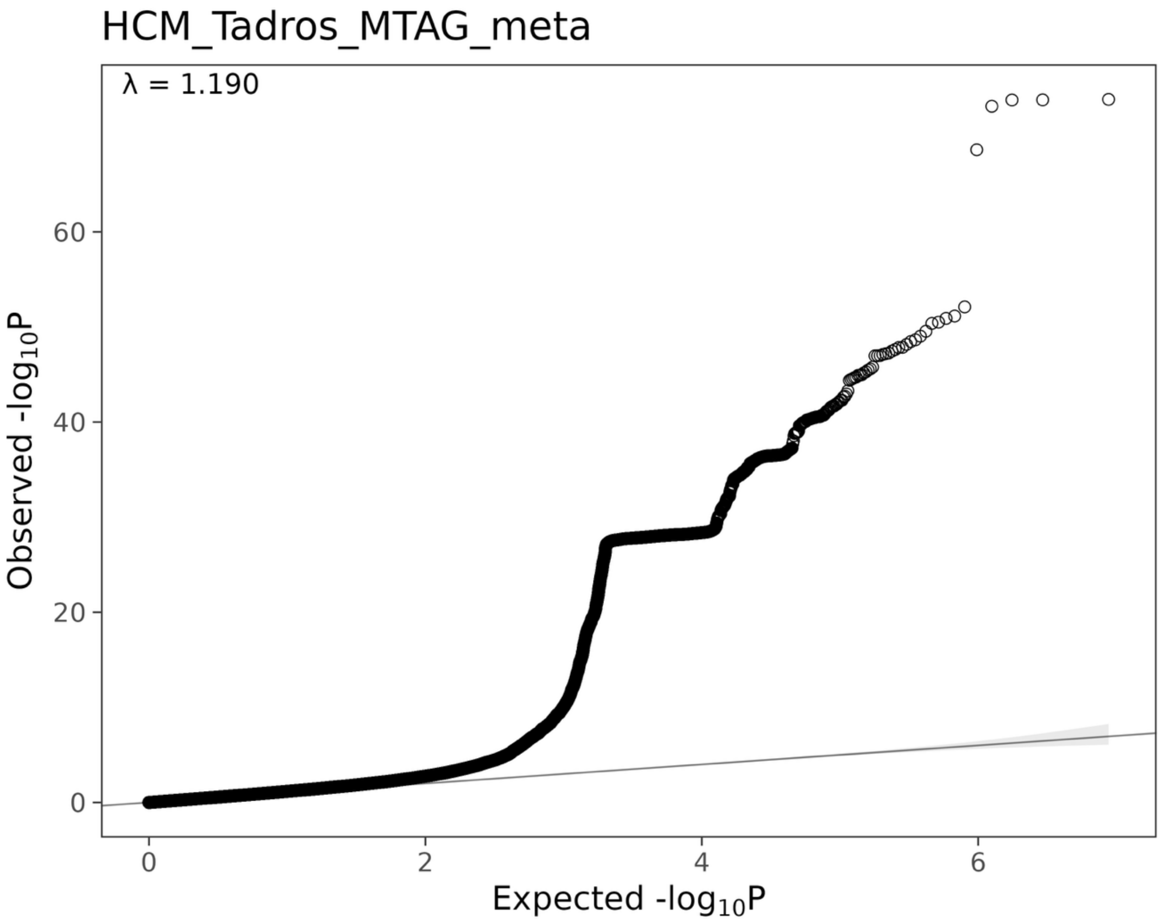
HCM_Tadros_MTAG_meta
λ = 1.190
Observed -log10P
Expected -log10P
0
20
40
60
0
2
4
6
